# Dreaming during Covid-19 pandemic: Computational assessment of dreams reveals mental suffering and fear of contagion

**DOI:** 10.1101/2020.05.19.20107078

**Authors:** Natália Bezerra Mota, Janaina Weissheimer, Marina Ribeiro, Mizziara de Paiva, Juliana D’Ávilla, Gabriela Simabucuru, Monica Frias Chaves, Lucas Cecchi, Jaime Cirne, Guillermo Cecchi, Cilene Rodrigues, Mauro Copelli, Sidarta Ribeiro

## Abstract

Neuroscience and psychology agree that dreaming helps to cope with negative emotions and learn from experience. The current global threat related to the COVID-19 pandemic led to widespread social isolation. Does dreaming change and/or reflect mental suffering? To address these questions, we applied natural language processing tools to study 239 dream reports from 67 individuals either before the Covid-19 outbreak or during March-April, 2020, when quarantine was imposed in Brazil following the pandemic announcement by the WHO. Pandemic dreams showed a higher proportion of anger and sadness words, and higher average semantic similarities to the terms “contamination” and “cleanness”. These features were associated with mental suffering related to social isolation, as they explained 39% of the variance in PANSS negative subscale (p=0.0092). These results corroborate the hypothesis that pandemic dreams reflect mental suffering, fear of contagion, and important changes in daily habits.

---

The relationship between dreams and fear has long been expressed across different cultures, since the dawn of civilization ^1^. What ancient cultures interpreted as divine advice has now been hypothesized as a memory mechanism to metabolize negative emotions and learn from experience, preparing individuals for the challenges in everyday life ^2^. We have been facing a global threat at the moment - the COVID-19 pandemic - and new strategies to overcome this danger have been proposed, such as social isolation. This affects people’s lives in different ways, from the reassessment of economic perspectives to the effort to maintain safe and meaningful relationships. How can dreams reflect our inner suffering and even mitigate or create solutions for our challenges?

There has been an intense academic debate about the function of dreams and the underlying brain mechanisms ^3,4^. One issue in such debate is how episodic memories (memories of lived experiences) are manifested during dreaming ^5-9^. A related question is whether threatening events - such as the pandemic spread of a virus - can influence people’s dreaming patterns. For individual trauma, the role of dreaming is already recognized, and recurrent dreams depicting threatening situations are an important sign in the diagnosis of post-traumatic stress disorder ^10-16^. However, the scenario is far less clear when it comes to collective trauma, such as the one we have been experiencing with COVID-19.

Like an episodic memory, the concept of ‘day-residue’, which became central to Freud’s psychoanalytic theory and therapy ^17^, refers to elements experienced during the previous days, which can be identified once dreams are carefully scrutinized. Although it seems to be irrefutable today that the waking life is part of dreaming, recent studies have shown that only isolated spatial and temporal fragments of episodic memory are present in dreams ^3,10,11^. However, when traumatic experiences are at stake, dream imagery can get highly episodic. One explanation is that when something out of the ordinary happens — like a pandemic — our brains may process the experience through dreaming. Episodic fragments (such as characters and settings), as well as semantic information, are brought together over time, maintaining a sense of first-person and continuous present perspective.

Dream reports collected after the 9/11 attacks show that many New Yorkers reported dreams of being overwhelmed by a tidal wave or being attacked and robbed ^12^. According to the authors of that study, dreams are thought to be the brain’s way of working out our emotional problems, and the more anxious we become in waking life, the more vivid our dream images seem to become.

Barrett et al. examined the dream contents produced by British officers held in a Nazi camp for prisoners of war (POW) between 1940–1942 ^13^. These POW dreams showed more content concerning battles, imprisonment, escape, and food than the male norms from the same era. Not much of any type of social interaction was detected in the POW dream reports, with less friendliness, sexuality, and even less aggression than the male norms. However, aggression was unusually extreme when it occurred, and its content was linked to previous battles rather than camp life. POW’s had less good fortune or misfortune in their dreams along with frequent bland dreams about the camp’s tedium.

Dream reports produced by German civilians during the Nazi regime were extremely bland, and the author’s main conclusion was that the oppressive environment was so extreme as to repress even dreams ^16^. One caveat is that most data about dreams collected during confinement or obtained from former war combatants have been gathered only after the person is away from the arena of combat ^12^. Studies of dream reports during extended confinement, like the present one, are scarce and called for.

The subjective nature of dreams challenges its scientific investigation, just like psychiatric evaluation. How to precisely characterize aspects from oral reports that in human contact we can describe with words, just like the specialist evaluation of human behavior in a psychiatric interview? To overcome this problem and allow an objective metric of content and structural analysis of oral reports, natural language processing tools have been applied with success in the field of psychometric evaluation ^18-25^. Structural analysis applying graph representation of word trajectory reveals mental disorganization related to negative symptoms in schizophrenia, allowing an automated diagnosis with more than 90% accuracy in different disorder stages and different languages ^19-21,24,26^.

Interestingly, dream reports seem to contain the most revealing content yielding this precise discrimination of schizophrenia diagnosis ^19,21^. The estimation of semantic similarity based on the co-occurrence of terms in a large corpus has enabled the identification of substance use and dose by calculating semantic similarity between experience oral reports and probe words related to psychotropic substances like methamphetamine ^23^. To conclude, estimation of the proportion of affective words with positive or negative valence has been useful to identify mood disorders such as bipolar disorder, also using dream reports ^27^. It also predicts the response of psilocybin treatment for resistant depression ^28^.

More than just tracking signs of mental suffering, could the observation of dreams help us understand personal challenges and mitigate suffering? Blagrove and collaborators proposed that dreams sharing has an empathic effect on the dreamer and on significant others engaged in a dream telling situation^29^. Trait empathy was found to be significantly associated with the frequency of listening to the dreams of others, frequency of telling one’s dreams to others, and attitude toward dreams. Moreover, the dream-discusser state of empathy towards the dream-sharer was found to increase significantly as a result of the dream discussion, with medium effect size, whereas the dream-sharer presented a small decrease in empathy towards the dream-discusser. Based on these results, the authors suggest that dreams act as a piece of fiction that can be explored by the dreamer together with other people, inducing, thus, empathy about the life circumstances of the dreamer. They also speculate that the story-like characteristics of adult human dreams may have been positively selected during human evolution as part of the selection for emotional intelligence, empathy, and social bonding.

Here we aim to use natural language processing tools to investigate the impact of the COVID-19 pandemic in dream experience, testing the following hypotheses: 1. The content and structure of pandemic dream reports are different from the same features obtained from dream reports collected before the outbreak of the COVID-19 virus and these peculiarities evolve; 2. The specific peculiarities of pandemic dreams are associated with mental suffering measured by psychometric scales, being 3. associated with the self-reported emotional impact of dream observation as well (Figure 1).

**Figure 1:**
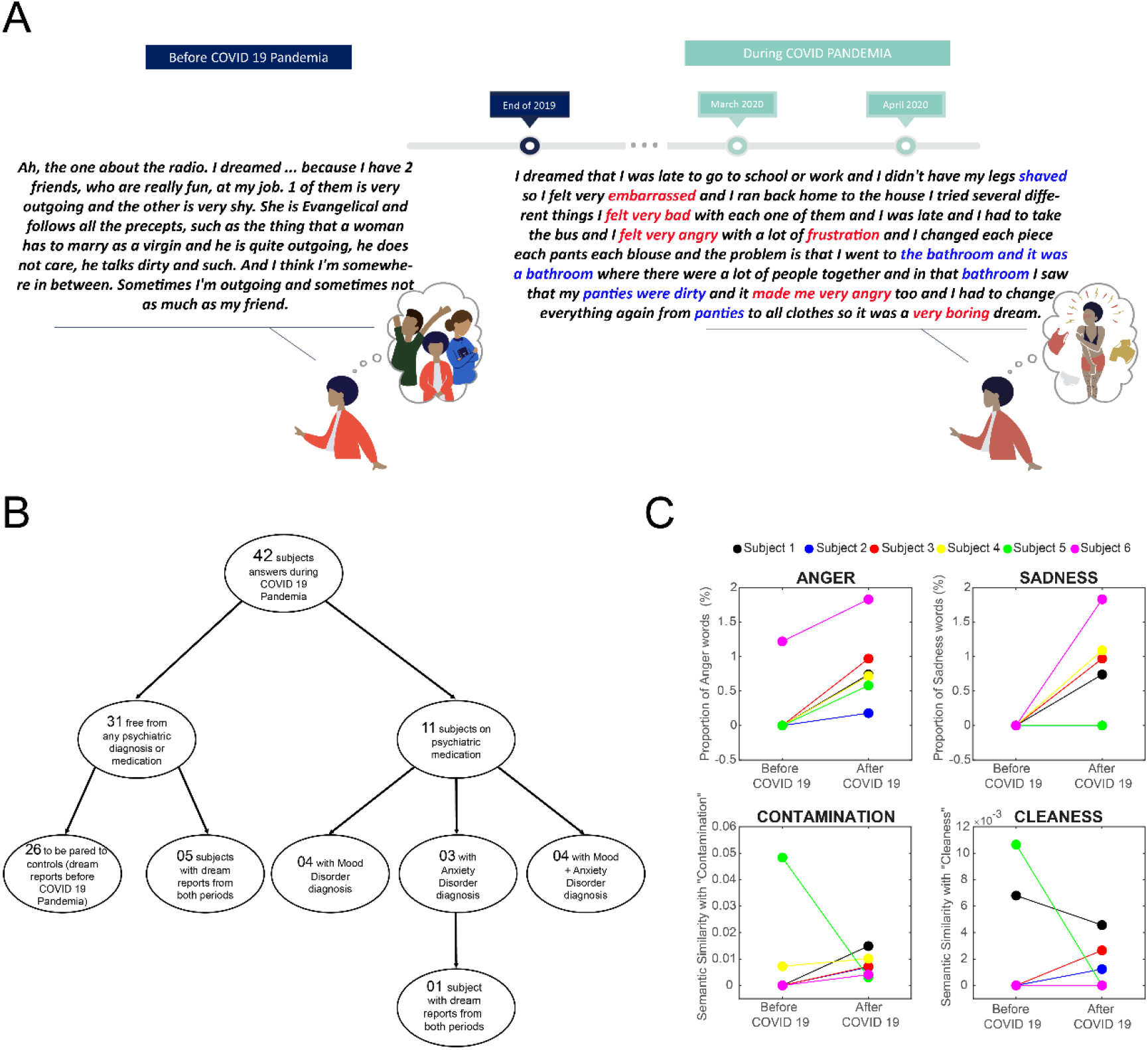
Are dream reports from the COVID-19 Pandemic period different from dream reports before the COVID-19 virus existence? (A) Timeline of data collection (audio recordings of dream reports through smartphone application), and illustrative examples of two dream reports from the same participant before and after the COVID-19 Pandemic. Red marks words linked to emotions and blue marks words semantically associated with the terms “contamination” and “cleanness”. (B) Fluxogram of group composition. (C) Representative examples of computational analysis of dream reports from the same participants before and after COVID-19 Pandemic.

## RESULTS

We followed 42 participants just after the pandemic announcement by the World Health Organization (WHO) and during the first month of social isolation in Brazil (March 12 – April 21), collecting oral dream reports through a smartphone application designed to gather audio data collection files for mental health monitoring. From this sample, 11 participants reported the use of psychotropic medication and mental health diagnoses related to mood and anxiety disorders (Figure 1B). Those participants were analyzed separately. A group of 31 healthy participants without mental health diagnosis and free from any psychotropic medication was analyzed. Five of them (and one participant that were using psychotropic medication during the pandemic period but were not before COVID-19) had already used the same smartphone application before the existence of COVID-19. These 6 participants plus 25 healthy participants that also used the smartphone application before the COVID-19 pandemic, composed a paired control group to be compared with the pandemic group (Figure 1B). These six participants’ dream reports are presented as illustrative examples for content analysis in Figure 1C.

### Comparison between Pandemic x Control Dreams and the longitudinal observation

The structural analysis did not differ between pandemic dreams and controls: despite pandemic dreams presenting a higher amount of words in general, no difference was found regarding word connectedness (Figure 2A, Table 2).

**Figure 2:**
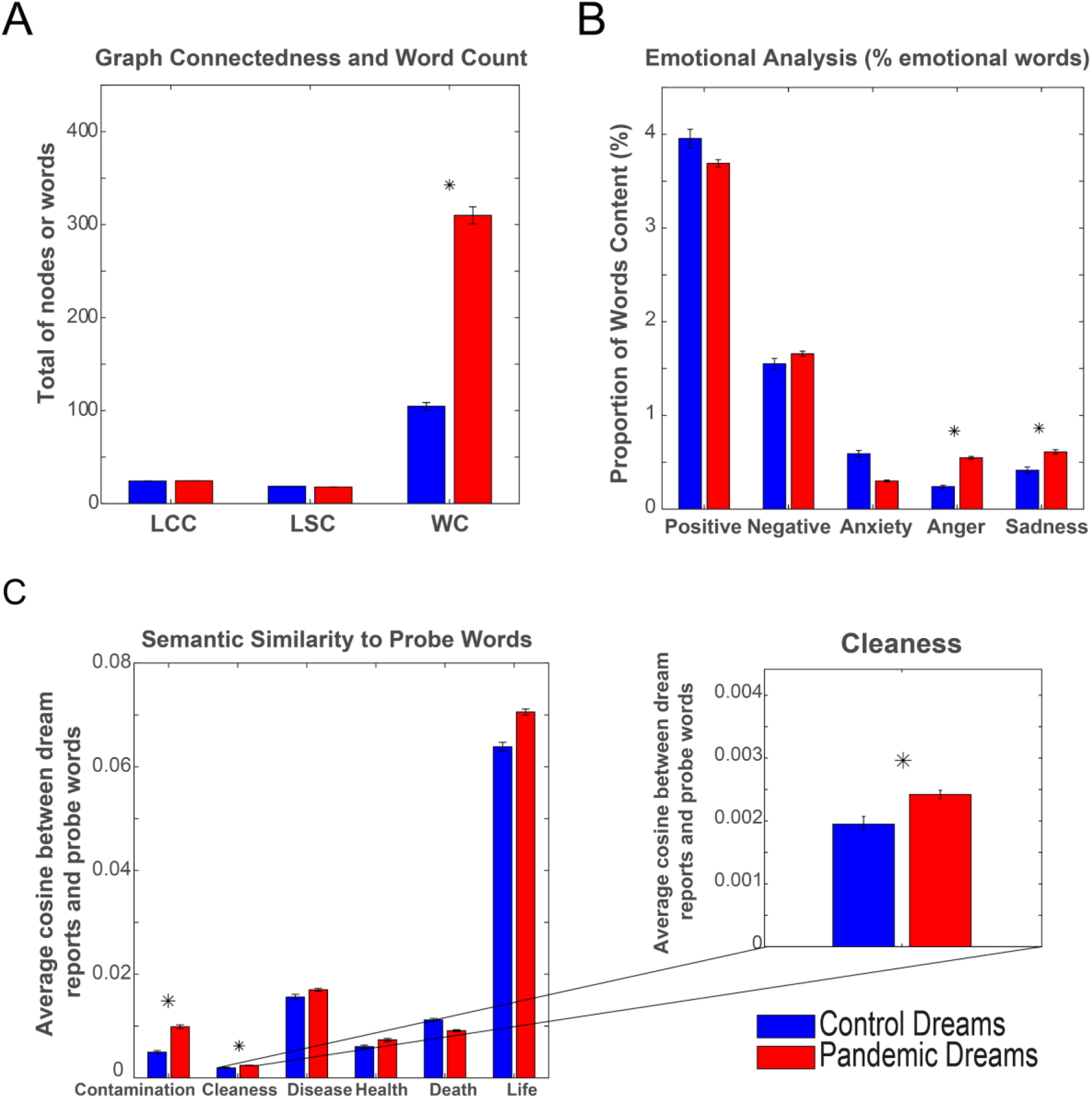
Pandemic dreams in comparison to control dreams (collected before COVID-19 Pandemia) (A) Structural analysis based on the representation of word trajectory as a graph. No significant difference for word connectedness measured by LCC or LSC. Word count was different between the groups (p < 0.0001, pandemic dreams are longer). * represents significant differences between groups after Bonferroni correction for multiple comparisons (p < 0.0167). (B) The proportion of emotional words in dream reports. Pandemic dreams present a higher proportion of words related to Anger (p = 0.0001) and Sadness (p = 0.0003) compared to paired control dreams. * represents significant differences between groups after Bonferroni correction for multiple comparisons (p < 0.0100) (C) Semantic similarity to probe words. Pandemic dreams present higher semantic similarities to the terms “contamination” (p = 0.0008) and “cleanness” (p = 0.0022). * represents significant differences between groups after Bonferroni correction for multiple comparisons (p < 0.0083).

Although structure seems to be preserved, emotional and semantic content revealed peculiarities: the proportion of anger and sadness-related words was higher for pandemic dreams. Semantic similarities to the terms “contamination” and “cleanness” were also higher (Figure 2B and C, Table 2). No difference was found for the proportion of positive emotions, negative emotions, or anxiety-related words, neither for semantic similarities to the terms “sickness”, “health”, “death” or “life” (Figure 2B and C, Table 2).

The analyses of connectedness and word proportion are already controlled for word count (see Methods), but semantic similarities might be impacted by the higher amount of words in pandemic dreams. For that reason, we verified Spearman correlations between word count and semantic similarities and no correlation was significant (Table 2).

Consecutively, we verified whether the pandemic dreams peculiarities increase or decrease over time, during the dream observation period. To do so, Spearman correlations with the time of dream observation was performed. The semantic similarity to the term “cleanness” presented an increase over time (Table 02). However, Spearman correlation did not reach significance level after Bonferroni correction (Rho = 0.17, p = 0.0290, Table 02).

### Association with mental suffering

Pandemic dreams peculiarities such as the proportion of anger or sadness related words, or semantic similarities to “contamination” and “cleanness” were combined and correlated with the scores from PANSS (a psychometric scale that evaluate positive symptoms linked to psychosis such as hallucinations and delusions; negative symptoms linked to social isolation and cognitive impairment; and a general subscale linked to unspecific mental suffering such as mood and anxiety symptoms, using canonical regression (Figure 3). We found a significant correlation between the combination of dream content peculiarities and mental suffering (Figure 3A). PANSS negative subscale and semantic similarity to the term "cleanness" contributed with higher coefficients to the canonical correlation, and both co-varied in the same direction (the higher the semantic similarities to “cleanness” in the dream report, the higher the severity of negative subscales in PANSS (Figure 3B). This result was confirmed by multi-linear correlation between the combination of pandemic dreams peculiarities and PANSS negative subscale or mental suffering related to social isolation (R^2^ = 0.39, p=0.0092, Coefficients: 1.3 Anger proportion + (-0.043) Sadness proportion + 18 “Contamination” similarity + 984.79 “Cleanness” similarity + 7.7). In other words, pandemic dreams peculiarities explained 39% of PANSS negative subscale variation. To better understand which items were particularly related to the similarity to “cleanness”, we calculated the canonical correlation between all 7 items of PANSS negative subscale and the semantic similarity to the term “cleanness” (Figure 3C). We found that most of the variance of this correlation was due to the co-variance between the semantic similarity of dream reports to the term “cleanness” and the items N3 of PANSS negative subscale (Poor Rapport), with a smaller contribution in the same direction of the item N4 (Passive Social Withdrawal), and in the opposite direction of the item N6 (Lack of Spontaneity on Talking). In other words, the more similar to cleanness the dream report was, the higher the severity of poor rapport, passive social withdrawal, and higher spontaneity on talking during the interview.

**Figure 3:**
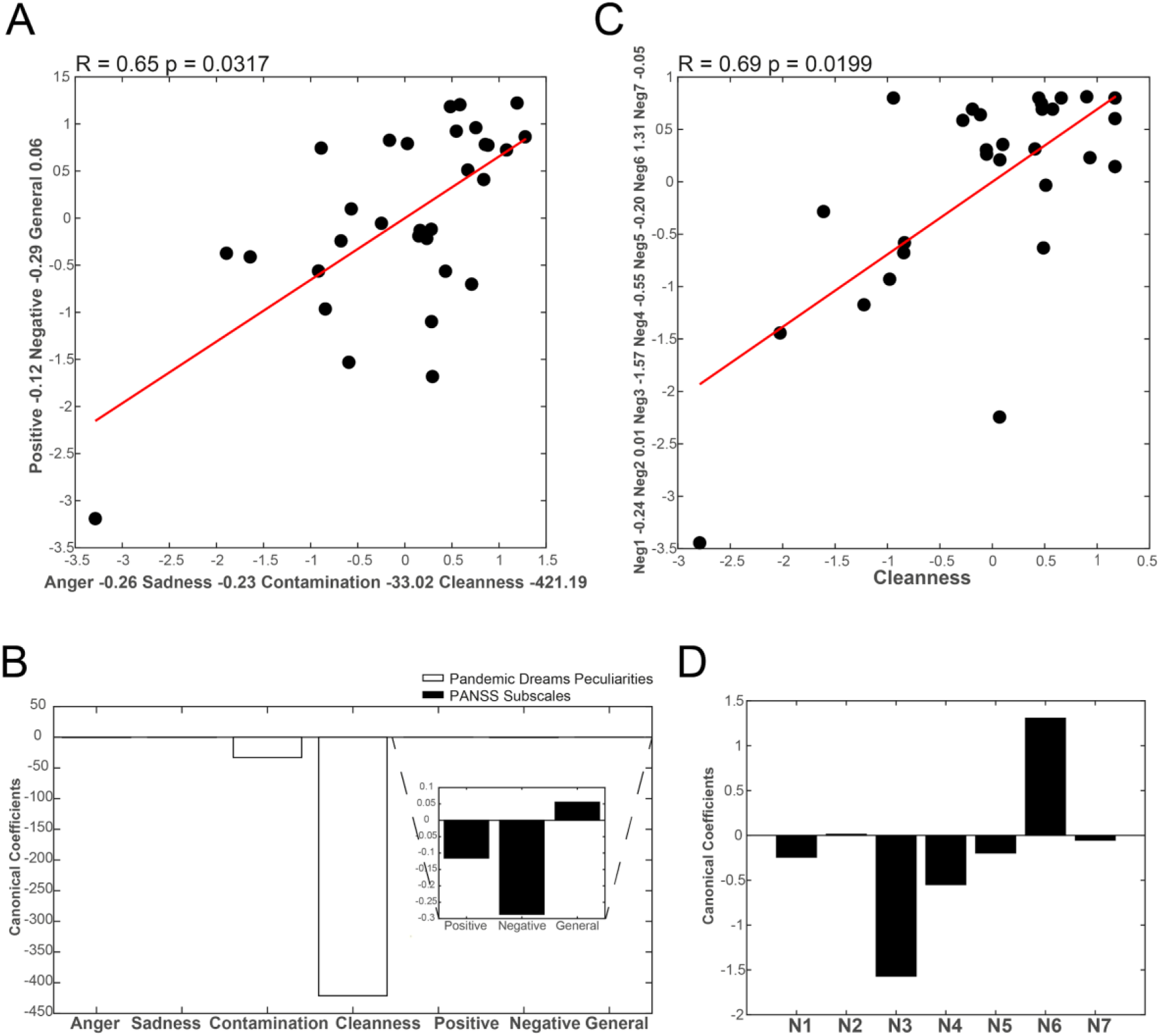
Canonical correlations of pandemic dreams peculiarities to psychometric evaluation (severity of mental suffering measured by PANSS) (A) Canonical correlations of pandemic dreams peculiarities (proportion of anger, sadness, and “contamination”, “cleanness” semantic similarities) to 3 PANSS subscales (Positive, Negative, and General symptoms). R and p-values on the title, and the canonical coefficients on the x and y axes. (B) Canonical coefficients of pandemic dreams peculiarities (white bars, showing the higher contribution of semantic similarity to the term “cleanness” to this correlation), and of PANSS subscale (black bars, showing the higher contribution of PANSS negative subscale). (C) Canonical correlations of semantic similarity to the term “cleanness” to 7 items of PANSS Negative subscales. R and p-value on the title, and the canonical coefficient on x and y-axis. (D). Canonical coefficients of PANSS Negative subscale (black bars, showing the higher contribution of PANSS negative subscale items N3 - Poor Rapport, N6 - Lack of spontaneity, N4 - Passive Social Withdrawal). The canonical coefficient of the semantic similarity to the term “cleanness” is -483.50.

We performed the same multilinear correlation analysis for a group of participants under psychotropic medication that self-reported mood and/or anxiety disorder. No significant correlation was found between peculiarities of pandemic dreams and psychometric evaluation, perhaps because the sample size was too small to reach a significant level (Total PANSS: R^2^ = 0.30, p=0.8458, Positive Subscale: R^2^ = 0.68, p=0.3675, Negative Subscale: R^2^ = 0.26, p=0.8843, General Subscale: R^2^ = 0.19, p=0.9369). Interestingly, however, the association with the positive subscale was the strongest found, a different pattern than for the group with no mental disorder diagnosis.

### Association with the affective impact of dream observation (self-evaluation scale)

To verify a potential affective impact of the dream observation process, at the end of the data collection we applied a self-evaluation questionnaire, in which participants rated, using a 1 to 10 scale, to which extent the dream observation experience increased their states of being happy/sad, calm/anxious, energetic/tired, peaceful/aggressive, altruistic/selfish, and creative/confused. We constituted pairs of positive/negative subjective evaluations. First, we compared the scores assigned, in order to verify how participants self-evaluated the dream-observation experience in positive and negative terms. We found higher positive evaluation for almost all the pairs of subjective impact analyzed (more happy than sad, more energetic than tired, more peaceful than aggressive, more altruistic than selfish, and more creative than confused) (Figure 4A). There was no difference between calm and anxious (Figure 4A). We then correlated all pandemic dreams peculiarities to the positive and the negative self-evaluated aspects and a significant canonical correlation to negative aspects of dream observation was found (Figure 4B). Also, while analyzing canonical coefficients, we noticed that most of the correlation variance is explained by the co-variance of the semantic similarity to the term “contamination” with the scores on the self-evaluation for aggressiveness (i.e., the more similar to contamination the semantic content of the dream report, the higher the feeling of being aggressive, or selfishness and self-confusion) (Figure 4C). However, the higher the similarity to “cleanness”, the lower the scores for aggressiveness, selfishness or self-confusion, and a higher effect of anxiety) (Figure 4C).

**Figure 4:**
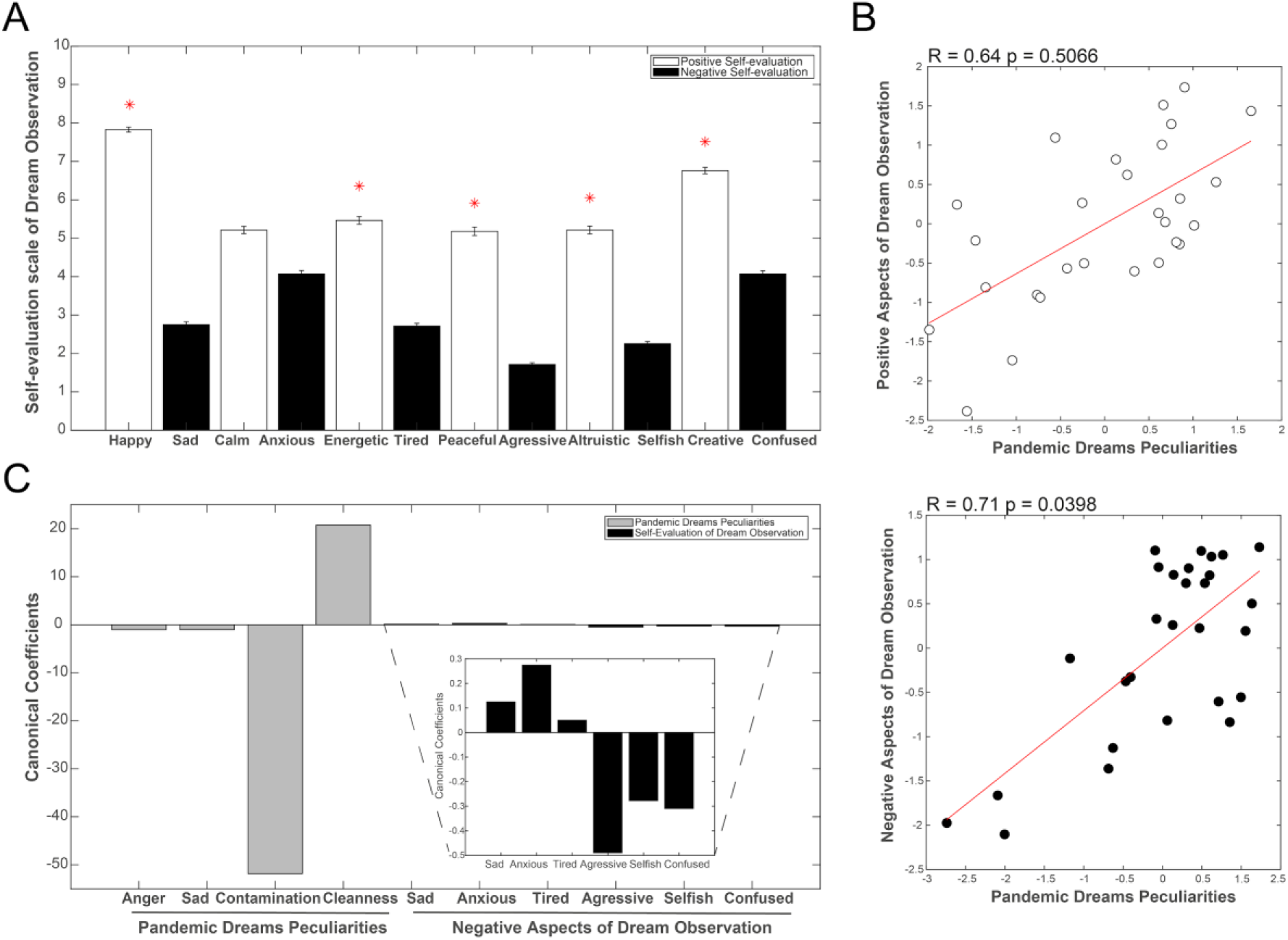
Self-evaluation of dream observation. (A) Comparison between positive and negative aspects of self-evaluation of dream experience. As positive aspects we investigated how the participants considered that the experience of dream observation during the first month of quarantine due to COVID-19 pandemic made them happy, calm, energetic, peaceful, altruistic and creative; in opposition to negative aspects: sad, anxious, tired, aggressive, selfish and confused (B) Canonical correlations between pandemic dreams peculiarities (Proportion of Anger and Sadness related words + Semantic similarities to the terms “contamination” and “cleanness”) and Positive (top) or negative (bottom) self-evaluated aspects of dream observation. R and p values presented on the title. (C) Canonical coefficients of pandemic dreams peculiarities (gray bars, showing the higher contribution of semantic similarity to the term “contamination” to this correlation), and negative aspects of dream observation (black bars).

## DISCUSSION

The results are in accord with the hypothesis that dreams from the pandemic period reflect negative emotions such as anger and sadness and similarity to “contamination” and “cleanness”. This seems to show in dream content all different sources of fear and frustration to this global challenge. Economic fears, social distancing from relatives, friends, and peers, and uncertainties on future planning are possible explanations for the above-reported changes on dream content and the increase of mental suffering.

Besides, pandemic dreams reflect changes in habits, more than worries related to greater impacts or tragic ends in people’s lives. There is a great similarity to daily concerns, with concrete changes in habits (related to contamination and cleanness), which tends to increase during the follow-up. Those are changes that are globally discussed and adopted ^30^. Nevertheless, disease and fear of death, during the first month of observation, were not concretely faced as a challenge for most of the participants (only one individual had confirmed the disease during the follow-up and another had a close relative with confirmed infection). Maybe this result could be different in another time point, or if sickness and death concretely appeared as a reality in sample majority. But what concretely affected the daily routine of individuals, the change of habits, and fear of contamination was tracked by the higher similarity to these terms.

Although this sample did not experience an individual severe trauma by fear of death or grief of losing a beloved person or a job, the change, in reality, was already enough for some individuals to experience mental suffering throughout this ongoing pandemic period. Moreover, the combination of dream content changes (higher proportion of anger and sadness combined with higher semantic similarity to “contamination” and “cleanness”) was significantly correlated with mental suffering related to aspects of social isolation, reflecting 39% of PANSS negative subscale variance. Even though this psychometric scale has been designed to quantify the severity of negative symptoms related to psychosis, the mental suffering associated with poor rapport and passive social withdraw, combined with more fluency while talking during the interview was associated with higher similarity to the term “cleanness”. Changes in dream content seem to reflect mental suffering similar to what happens with patients experiencing harsh changes in reality at a psychotic break. Not that this suffering was severe to the point of being considered pathological, but, as reality during a pandemic period seems too threatening, dream contents reflect this modulation, capturing this quality of suffering from the loss of social interaction.

Interestingly, a subsample of individuals with a diagnosis of mood and/or anxiety disorder did not show the same level of significant association with the same class of symptoms. Although the sample size of this group is too small to be conclusive, R-squares seem to indicate that there might be a real association, emphasizing the importance of increasing the sample size. Furthermore, psychotropic medications have a major impact on dreaming ^31,32^.

Importantly, changes in structural language analysis were not observed. Those changes have been associated with cognitive impairments in psychosis or during development, being, thus, a deep effect, not expected to be found in scenarios like the one under analysis. Also, during the pandemic period, dream reports were longer, wordier. This may indicate that humans are paying more attention to dream experiences at the current moment. It might as well suggest an increase in the ability to remember dreams details due to the possibility of staying in bed after waking, during the quarantine, although this factor did not interact with other results (correlations were not significant).

Moreover, observing dreams were more positively evaluated. This result may indicate a relatively safe way for self-observation and mental health management that can be oriented during this period of reality uncertainties ^29^. The simple act of observing and reporting dreams can have a positive impact on mental health. Also, dream reports peculiarities can signal the need to be cautious: on one hand, the more similar dreams were to the term “contamination”, the more people reported being aggressive, selfish, or confused after dream observation. On the other hand, the more similar the dreams were to the term “cleanness”, the higher the feeling of being anxious after dream observation. Dream observation seems, thus, informative about mental health issues, showing different perspectives of self-evaluation.

This study has several limitations such as a small sample size, without representative diversity related to gender balance, age, or educational level. Although this might impede generalizing the obtained result to the global population, it does suggest a relatively simple and safe manner to track mental suffering during periods of forced social isolation. Larger studies in diverse samples should light our understanding of dreams as a reflector of inner suffering and fears. It might as well help us identify early signs of mental issues, being thus part of the prevention of severe mental impact.

In conclusion, our results support the theory that dreams reflect daily changes during this critical period of our lives, associated with mental suffering. Observing and reporting dreams, thus, may help individuals to overcome this challenge, flagging feelings such as anxiety and sadness.

## METHODS

### Participants

To address the 3 hypotheses formulated we followed 42 participants during the initials days of lockdown in Brazil after the announcement of COVID-19 pandemic by the World Health Organization WHO (from March 12th to April 21st), according to each participant location (Figure 1B and Table 1). As eleven of them reported continuous use of psychotropic medication and mental health diagnosis, we analyzed their data separately (Figure 1B). Then we constituted a control group of 31 participants pared to the 31 participants free from mental health diagnosis (Figure 1B and Table 1). All the control participants were also free from mental health symptoms, diagnosis, or psychotropic medication. They used the same method to collect dream reports remotely before the existence of the COVID-19 virus (using a customized smartphone application designed for access to memory oral reports to track for mental health suffering signs). Six of the 31 participants of the control group could also participate in this pandemic period of data collection (Figure 1B), and these 6 paired examples of changes in dream reports are shown in Figure 1C. Mean and standard deviation of age and educational level are summarized in Table 1, as well as the sex distribution. All the participants agreed to participate and signed informed consent. The study was approved by the UFRN Research Ethics Committee (permit # 3.613.186).

**Table 1:** The Pandemic and Control groups were balanced for years of age and formal education, as well as sex. Age and education show mean, standard deviation, and p-value for Wilcoxon ranksum comparisons. Sex shows the number of males (M) and females (F) and p-value after Chi-square comparison between groups.

|  | Age (years) | Education (years) | Sex |
| --- | --- | --- | --- |
| **Pandemic dreams group** | 35.23 ± 8.72 | 15.50 ± 2.39 | 5M/26F |
| **Control dreams group** | 32.57 ± 9.13 | 15.35 ± 1.50 | 6M/25F |
| **Pandemic x Control (p value)** | 0.0665 | 0.4019 | 0.7396 |

**Table 2:** Comparison of Pandemic and control dream reports and correlations with verbosity (measured by total word count) Comparisons using non-parametric Wilcoxon Ranksum test Bonferroni corrected (p-value threshold indicated for each analysis). * indicates results considered significant. The significant differences were used to choose the features to be correlated with word count and also correlated with time of observation (measured in days after started the lockdown).

| Wilcoxon Ranksum Comparison Pandemic x Control Dreams |  |  |  |  |  |  |
| --- | --- | --- | --- | --- | --- | --- |
| Semantic Similarities<br>p (* < 0.0083) | contamination<br>0.0008* | cleanness<br>0.0022* | sickness<br>0.1467 | health<br>0.0589 | death<br>0.2413 | life<br>0.0738 |
| Emotional content %<br>p (* < 0.0100) | positive<br>0.7408 | negative<br>0.1598 | anxiety<br>0.1069 | anger<br>0.0001* | sadness<br>0.0003* |  |
| Structural Analysis<br>p (* < 0.0167) | LCC<br>0.2975 | LSC<br>0.0859 | WC<br>0.0000* |  |  |  |
| Spearman Correlation to WC |  |  |  |  |  |  |
| Pandemic Dreams peculiarities<br>p (* < 0.0125)<br>Rho | anger<br>0.1367<br>-0.27 | sadness<br>0.4164<br>0.15 | contamination<br>0.3836<br>-0.16 | cleanness<br>0.2085<br>0.23 |  |  |
| Spearman Correlation to time of dream observation (days after started the lockdown) |  |  |  |  |  |  |
| Pandemic Dreams peculiarities<br>p (* < 0.0100)<br>Rho | anger<br>0.2323<br>0.10 | sadness<br>0.6336<br>-0.04 | WC<br>0.6151<br>0.04 | contamination<br>0.5189<br>0.05 | cleanness<br>0.0291<br>0.17 |  |

### Protocol

Before the Pandemic period, the participants were invited to test a smartphone application designed to remotely collect oral memory reports to be used for remote mental health tracking during September to November 2019. The application prompted a request to report a dream that had to have at least 30 seconds to keep the flow of memory reports. In sequence, the application requested a report from the previous day and prompted 3 images to then request to tell a story based on that picture. All the audio files were already and safely transferred to the university server and researchers transcribed all the audios into text files. For this study, we focused on the dream reports memory, to compare it to pandemic dream reports. The other memory reports were not considered. A total of 31 dream reports from this period well before the COVID-19 Pandemic were generated.

During the Pandemic period, the participants had to initially answer an online survey related to general information like sex, age, educational level, family income, sleep pattern, and mental health diagnosis and/or medication in use. They were instructed to daily record their dreams in an audio file (using the smartphone application designed for scientific purposes or other online resources that were available) and share it with the researcher to transcribe it to text files. A total of 208 dream reports from this period (first month of COVID-19 Pandemic) were generated and averaged per subject to compare with pared controls.

At the end of this observation period, a trained psychiatrist applied the PANSS scale to measure the severity of mental suffering related to this pandemic period. This scale is divided into 3 subscales and the major focus is on psychosis. The first sub-scale is composed by 7 questions and called as the positive symptoms scale, that focus on symptoms such as delusions and hallucinations; the second is also composed by 7 questions and is called the negative sub-scale, and related to symptoms associated with social withdraw, isolation, cognitive impairment, and recursive thinking. And the last sub-scale is composed of 16 general questions related to mood and anxiety symptoms such as tension, depression, panic attacks, time/space orientation, or lack of judgment. All questions are graded from 1 to 7.

To conclude, we applied a final online survey related to the self-evaluation of the subjective impact of dream observation during this period. The scale requested the participants to grade from 1 to 10 to how the dream observation experience made them: happy, sad, anxious, calm, energetic, tired, peaceful, aggressive, altruistic, selfish, creative, or confuse.

### Text Analysis

After the transcription of all the words on dream reports, we performed 3 different computational language analysis using different strategies:

Structural Analysis using SpeechGraphs software ^20^: for this analysis, each word in the text file was considered as a node and the temporal sequence between words were represented as directed edges. Here we focused on the total word count (WC) and connectedness attributes (LCC – the number of nodes in the largest connected component and LSC – the number of nodes in the largest strongly connected component) as these attributes were the ones most associated with mental health disorder (such as Schizophrenia ^19-21,24^). As there was no maximum limit for an oral report of a dream, to control for word count differences, we performed graphs of 30 words, using a step of one word to perform the next graph ^20^. Then, we calculate the LCC and LSC of all 30 word-graphs and average all LCCs and LSCs from the same reports.

The proportion of words using LIWC with Brazilian Portuguese Dictionary: we used the LIWC software ^33^ that searches and classify words in the dream reports according to a validated Brazilian Portuguese Dictionary ^34^ to find words with emotional content (positive emotions, negative emotions, anxiety, anger, and sadness related words). The software returns the proportion of words with the specific content in the dream report. As it is already a proportion, the result is controlled by word count differences.

Semantic similarity using customized software to perform Fast text method in Python: In order to investigate how the dream content must reflect pandemic related concerns or change of daily habits, we estimated the semantic similarity of dream reports to the probe words (“contamination”, “cleanness”, “sickness”, “health”, “death”, “life”). We did this using word embedding techniques. Word embedding is based on the *distributional hypothesis*, which states that semantically similar words tend to appear in similar contexts ^35^. Given that the reports were in Portuguese, we used the *Fast text* pre-trained multilingual word embedding ^36,37^. This model is built using a sliding window of predefined length that moves along a Portuguese Wikipedia corpus. At each step, a neural network is trained to predict the central word from the context, that is, from the other words in the window. Once the neural network is trained, a vector representation for each word in the corpus is extracted from the input and output layers of the model, which is used as the word embedding. The original text files were transformed to lowercase, word-tokenized, and cleared from non-alphabetic tokens and stop-words (using nltk Portuguese stop-word list ^38^). Then, for each text file, we calculated the semantic similarity of each word to the probe words by calculating the cosine between the word and the probe word. To control for the possible difference in word count and also to refine the results for only the most related words to the probe terms, we considered to the final average similarity only those words that reached the threshold of 0.3 (the maximum similarity is 1). All the words that reached at least this threshold were considered and averaged to compose a final semantic similarity of the dream report to the probe words.

So, the semantic similarity of a dream report to a probe word was calculated as the average similarity of all the words on dream reports that reached the minimum similarity of 0.3 to the probe word. Semantic similarity between a word (***v_1_***) to a probe word (***v_2_***) is the cosine between these two vectors:

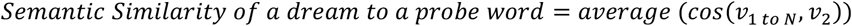

where |**v_2_**| refers to the probe words (“contamination”, “cleanness”, “sickness”, “health”, “death”, “life”) and |**v_1_**_to N_| refers to the all the words on dream reports that presented at least a similarity of 0.3 to the probe word considered.

### Statistical analysis

To address the first hypothesis, we compared the dream reports produced during the COVID-19 pandemic period (Pandemic Dreams) to dream reports produced before the existence of the COVID-19 virus. For that, we average all dream reports from the same participant produced during the pandemic period, to have only one data point for each participant. As the data was not normally distributed (all the text analyses were tested for normality distribution using the Kolmogorov Smirnoff test and all p values were lower than 0.0001), we used a nonparametric test Wilcoxon Ranksum. We stratified the analysis as structural analysis (3 attributes: WC, LCC, and LSC), the proportion of emotional content analysis (5 attributes: positive emotions, negative emotions, anxiety, anger, and sadness related words) and semantic similarity analysis to 6 probe words (“contamination”, “cleanness”, “sickness”, “health”, “death”, “life”). We corrected for multiple comparisons considering each analysis, so we considered significant results of p-value lower than 0.0167 for structural analysis, 0.0100 for the proportion of emotional analysis, and lower than 0.0083 for semantic similarity to probe words.

For the longitudinal analysis, we correlated only the dream peculiarities found on the previous comparison and correlated to the time of dream observation counted in days. As we found 5 attributes, we corrected for multiple comparisons using the Bonferroni method and considered difference p values lower than 0.0100.

To investigate the association to mental suffering, we combined and correlated all the pandemic dream content differences found to the PANSS sub-scales (positive, negative and general) using canonical correlation, and confirmed the result by performing a multi-linear correlation between the PANSS subscale with the higher canonical coefficient with the pandemic dreams variables. To better understand which symptoms were leading the correlations, we performed a canonical correlation between all questions in the PANSS subscale and the pandemic dreams variable that showed the higher canonical coefficient. To study the group of individuals with a psychiatric diagnosis and under medication, we performed only the multi-linear correlation (to avoid overfitting combining multiple variables as this group was too small).

To investigate how the experience of dream observation was self-evaluated with the dream reports peculiarities, we first compared 6 positive aspects of this questionnaire with 6 paired negative aspects of this scale, correcting the results for multiple comparisons using Bonferroni method (considering significant p < 0.0083). In sequence, we correlated the positive and the negative aspects with pandemic dreams peculiarities found using canonical correlation and studied the coefficients from the two sets of variables (to understand those that co-varied).

## Data Availability

All the data are available under request.

## Acknowledgments

We want to thank the financial support from Boehringer-Ingelheim (grants FADE/UFPE 270,906 and 270,561), Conselho Nacional de Desenvolvimento Científico e Tecnológico (CNPq) PVE grant 401518/2014-0, Universal grants 480053/2013-8, 408145/2016-1, 439434/2018-1, 425329/2018-6 and Research Productivity grants 308775/2015-5, 306659/2019-0, 301744/2018-1 and 310712/2014-9; Coordenação de Aperfeiçoamento de Pessoal de Nível Superior (CAPES) Projects OBEDUC-ACERTA 0898/2013, PROEX 534/2018 and STIC AmSud 062/2015; Fundação de Amparo à Ciência e Tecnologia do Estado de Pernambuco (FACEPE) grant APQ--0642--1.05/18; Center for Neuromathematics of the São Paulo Research Foundation FAPESP (grant 2013/07699-0). We also want to thank Ana Luiza Medeiros for illustrations in Figure 1 and all the participants.

## Author contributions

NBM designed the project, developed the smartphone application, collected the data, analyzed the data, and wrote the paper. JW pre-processed the data and contributed to writing the paper. MR, GS, and MC collected and pre-processed data. MP and JA pre-processed the data. LC and JC developed and coded the smartphone application. GC supervised smartphone application development. CR, MC, and SR supervised data collection, analysis, and contributed to paper writing.

## Materials & Correspondence

Correspondence and material requests should be addressed to the corresponding and first author NBM (Natália Bezerra Mota).

## Data availability

All the data are available in Supplementary Information.

